# Covid19, 2020 - a tutorial of sorts on reading data

**DOI:** 10.1101/2020.07.12.20151860

**Authors:** Isaac Meilijson

## Abstract

The SIR differential equations in Epidemiology are re-examined in the context of Covid19, 2020. The number of recovered cases is calibrated in time. Methods for estimating all pertinent parameters are described. A notion of implied susceptible population size ISPS is introduced, as the potential target population size for which the solution to the SIR equations would yield the current number of new affected cases. Analysis is applied to the Covid19 2020 data of a number of countries, as reported by the Johns Hopkins University data repository.

## 1 Introduction

The SIR (Susceptibles, Infected, Removed) model introduced by Kermack & McKendrick in 1927 [7] for the progress of an epidemic describes the interdependence between the cumulative number *X*(*t*) of affected cases, the cumulative number *R*(*t*) of removed cases (dead or recovered) and the ensuing current number of infected cases *Y* (*t*)= *X*(*t*) − *R*(*t*). Henceforth, the words “cumulative” and “currently” will be tacitly intended. The version of this model to be applied in the current report conforms with the usual assumption (henceforth the SIR “*γ*-equation”)

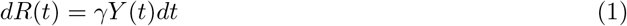

that the number of new removed cases constitutes a fixed proportion of the infected cases, and the usual assumption that the number of new affected cases is proportional to the product of the number *K* − *X*(*t*) of susceptible cases and an increasing function of the number of infected cases. However, *K* is fitted from data instead of letting it be the total population size, and the second factor (commonly modelled as the identity function) is taken (after Grenfell, Bjørnstad & Filkenstädt [4] to be a fractional power *Y* (*t*)*^α^*(0: *α<* 1) of the number of infected cases. In other words (the SIR “*β*-equation”)

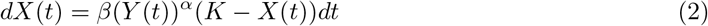

It is common practice to view *β* as varying with time, and watch closely for the date at which the effective reproduction rate *R*_0_, the ratio of *Kβ* to the removal rate *γ*, crosses from above to below 1. Thus, equation (2) (with *Y* instead of *Y^α^* and *K* as total population size) is perceived as a local relation, and not as a differential equation to be solved. We intend to establish via Covid-19 data, that the introduction of a power function of *Y* and of an adaptive target population size *K* make equation (2) solvable with constant *β*.

A companion publication by Alon and Meilijson [1] describes (*i*) methods to estimate the parameters *α*, *β*, *γ* and *K b*y maximum likelihood estimation under diffusion models based on the SIR differential equations, and (*ii*) a regression pre-processing procedure that modifies minimally the infected and removed components of the empirical data (*X*_1_*, Y*_1_*, R*_1_)*,⋯*, (*X_n_, Y_n_, R_n_*), so that the proportionality of new removals and infected cases will better hold. Item (*i*), that ignores the SIR *β*-equation (2), is expanded and modified in Section 2. Item (*ii*) is analyzed anew in Section 4 with simpler regular regression methods.

The data analyzed in this report is taken from the COVID-19 Data Repository of the Center for Systems Science and Engineering (CSSE) at Johns Hopkins University, from January 22, 2020 to August 2020. The purpose is to propose techniques to read and interpret these data, and to a lesser extent to build scenarios to project values into the near future. Attempts are made to take into account warnings and guidelines, such as those of Holmdahl and Buckee [6].

The (country-dependent) training period considered stationary extends roughly from the beginning of March to sometime between late May and early July. The working paradigm is that the three proportionality parameters *α, β, γ* estimated in the training period, stay in effect for the particular epidemic beyond the training period. The validity of this paradigm regarding *γ* will become apparent in Section 2. The model as a whole is not strongly identifiable: once the other parameters are adjusted, a relatively wide range of *α* values fits data adequately well, and perhaps one compromise value could fit all countries, but the analysis will be done per country. Validity regarding *β* can and will be assessed only in stationary periods where *K* is kept fixed. Under constancy of *β*, all variability will be phrased in terms of *K*, as follows. Equation (2) is rewritten as

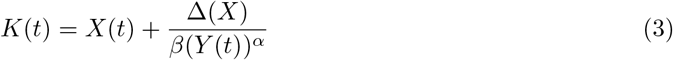

to be interpreted as the *implied susceptible population size* ISPS. Letting ∆(*X*) be a smooth version of *X*(*t* + 1) − *X*(*t*), *K*(*t*) is evaluated daily to provide an idea of the current target population of the epidemic. Due to the persistent missing data on weekends, the smooth version is taken throughout to be a moving average on the week around the date in question.

The notion of implied susceptible population size ISPS mimics implied volatility in Finance. The Black & Scholes ([3]) formula for option pricing does not quite hold when the asset estimated volatility is substituted, but it is such a convenient language tool that reverse engineering as in (3) is applied, and implied volatility is defined as the value for which formula and empirical price are the same. This value changes with the maturity date and the moneyness, and this non-constant shape is termed “volatility smile”. This is a word we will not export from Finance into Epidemiology.

ISPS is analyzed in Section 6 and illustrated in Figure 5 on six countries, projecting into the near future the solution to the SIR equations, with *β, γ* and *α* as fitted in the training period.

A word on the parameter *α*.

Imagine a small-world network topology composed of sparsely connected islands that are internally densely connected. Once an island is affected, contagion will apply locally. Much as in the Harris contact process [5], if the affected cases are considered a growing ball that transmits the disease by its perimeter, 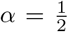 models the intra-island spread force. The “ball” could have some fractal dimension rather than a solid area. Alon and Meilijson [1] present an argument that on infinite populations, *α* = 1 yields exponential growth of *X*, while *α<* 1 gives rise to linear growth of *X*, with *Y* (*t*) increasing asymptotically to oscillate around the finite constant 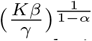. Formula (2) explains why should the proper parameter be *Kβ* rather than *β*, for a large population.

The average daily number of new affected and removed cases would then be 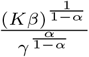. These levels, to be reported for every country under study, are so high that it is reasonable to infer that only a fraction of the population is susceptible.

Without claiming to delve deeply into epidemiology, it is suggested that isolation measures achieve stationarity by cutting contagion across islands. Relaxing isolation measures may have the effect of allowing for more contagion between islands, and this is what ISPS is intended to reflect.

Israel went through a Covid19 first wave that seemed to saturate, but relaxation of measures brought a second wave. USA is treated as one unit (perhaps wrongly, but state-by-state data is not detailed enough), and as such never quite stabilized. Unlike Israel and USA, the neighboring European countries Germany, Italy and Switzerland seem to have maintained stability at least until mid July, by a combination of official isolation measures and personal responsibility that has prevented for some time a return to the pandemic core. Figure 4 shows the dates considered stable in each of these countries. The epidemic is still growing in Brazil in mid August.

The purpose of this report is to show a technique for parameter estimation and recovery and infection calibration, and to suggest possible uses of ISPS as a measure of growth and perhaps as a driving wheel of epidemic management, without taking sides on discussions about management. With this said, under the strong evidence that *α<* 1, the author support the postulate that Covid19 grows in homogeneous sub-populations slower than exponential. However, increasing ISPS may indicate infection into new horizons, calling for isolation measures.

**A word on data quality**. Each country is analyzed on its own sake. The rate *γ* at which infected cases are removed should be (by the SIR *γ*-equation (1)) the slope of the linear regression of the newly removed cases on the number of infected cases. The plots of these linear regressions in Figure 2 are reasonable, but slopes *γ* vary too much from country to country. It is worthwhile to point out in this context that data is taken at face value. Although analysis is based on affected and infected cases only, the proportion of dead cases among removed cases will be evaluated and displayed in Figure 6. Israel and Europe data show this proportion to oscillate around a few percentage points and stabilize in the last two months of monitoring between 0.5% and 1%, with daily removal rate *γ* between 4% and 10%. In contrast, USA data shows lethal proportion oscillating around 50% and stabilizing at 3.5%, with daily removal rate *γ* below 1%. Brazil stands somewhere in between. There seem to be big differences between countries on what fraction of what data is actually reported, reported, with some countries under-reporting the number of recovered cases.

## 2 Removed cases -estimation of I and calibration of recovered cases delay

*This Section hinges on the common observation that recovered cases are reported later than dead or affected cases. As a result, the common naive subtraction operation that assesses the current number of infected cases, is positively biased. A method will be presented to gradually shift recovered cases backwards in time to the point that the SIR γ-equation is properly satisfied. At this point the number of infected cases will be minimally perturbed so that the SIR γ-equation is perfectly satisfied. This will provide an assessed day by day number of infected and removed cases, and an estimate of γ. The balance between dead and recovered cases will be ignored along the procedure, but monitored on the end result. The output of this procedure, with the original empirical cumulative number of affected and dead cases and the modified number of recovered cases, is the input for the next Section, dealing with the SIR β-equation: estimation of α and β via flexible K*.

The clear-cut SIR equation *dR*(*t*)= *γY* (*t*)*dt* should be thought of empirically, towards the formulation of regression equations, as

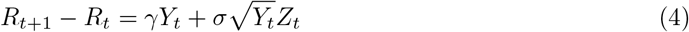

where *Z_t_* are ideally independent with mean zero and unit variance. Think conceptually of *R_t_*_+1_ −*R_t_* as binomially distributed with number of trials *Y_t_* and probability of success *γ*.

In other words, a homoscedastic regression model would express (4) as

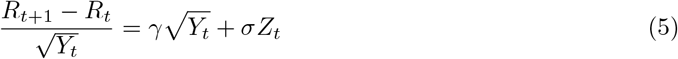

from which the regular Least Squares estimate of *γ* is

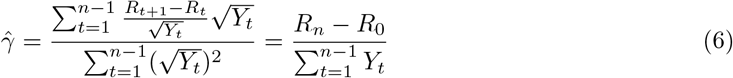

Exact solution *Ŷ* to the SIR *γ*-equation.

The above idea towards estimation is the starting point of the proposed pre-processing step. Recalling that *R* = *X* − *Y*, a perfect match of the SIR *γ*-equation would entail

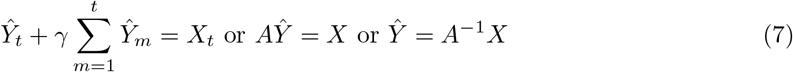

where *A* = *I* + *γB* and *B* is the lower triangular matrix with zeros above the diagonal and ones on and below the diagonal.

Empirical data are noisy. Reported affected and removed cases are at best a rough description of reality. It is proposed to respect the reported affected and dead cases but adopt a solution *Ŷ* of equation (7) as the number of infected cases. The parameter *γ* will be estimated by the value for 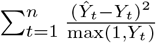which is minimal. I.e., *Ŷ* is the minimal tampering with the number of infected cases that will satisfy the SIR *γ*-equation.

**Figure 1:**
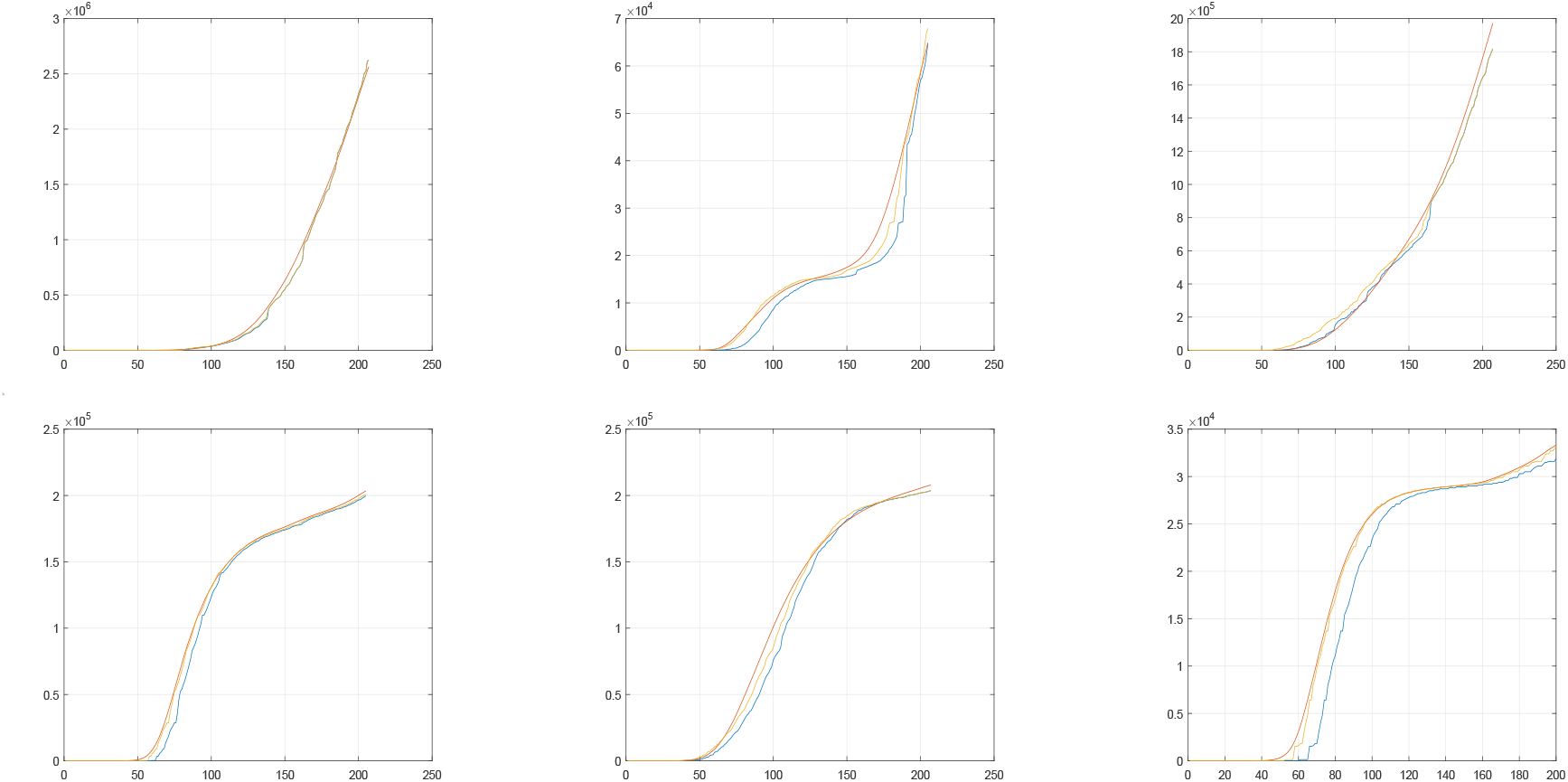
Number of recovered cases. Blue: raw data, yellow: shifted raw data, red (working version): inferred from regression-adjusted infected cases. From top left to bottom right: Brazil, Israel, USA, Germany, Italy and Switzerland.

**DAY -late report of recovered cases**. Actually, the method as such is inappropriate. A plot of *Y* and *Ŷ* reveals that the two peak at different times. The report of recovered cases seems to be delayed more than the other statistics. In the absence of a model, the parsimonious initial working paradigm is that this additional delay is a constant number of days *DαY*. In other words, the effective number of removed cases 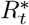 until date *t* is to be taken as the number of dead cases until date *t* plus the number of recovered cases until date *t* + *DAY*. The effective number of infected cases 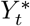 at date *t* is then 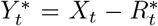. The parameters 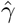 and *DAY* are jointly determined by minimizing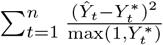.

Figure 1 illustrates the results of this method on each of the six countries. The Brazil and USA data fits well daily with no delay, and the other four manifest delays between 5 and 10 days. It is possible to see that the recovered-cases shifted empirical data *Y*^*^and the regression-fitted *Ŷ* are close to each other (somewhat less so in Italy) and rather far from the unshifted empirical data *Y*. That is, calibration of the recovered cases by the delay DAY, brings the (*S, I, R*) data to satisfy the SIR *γ*-equation more accurately.

**Figure 2:**
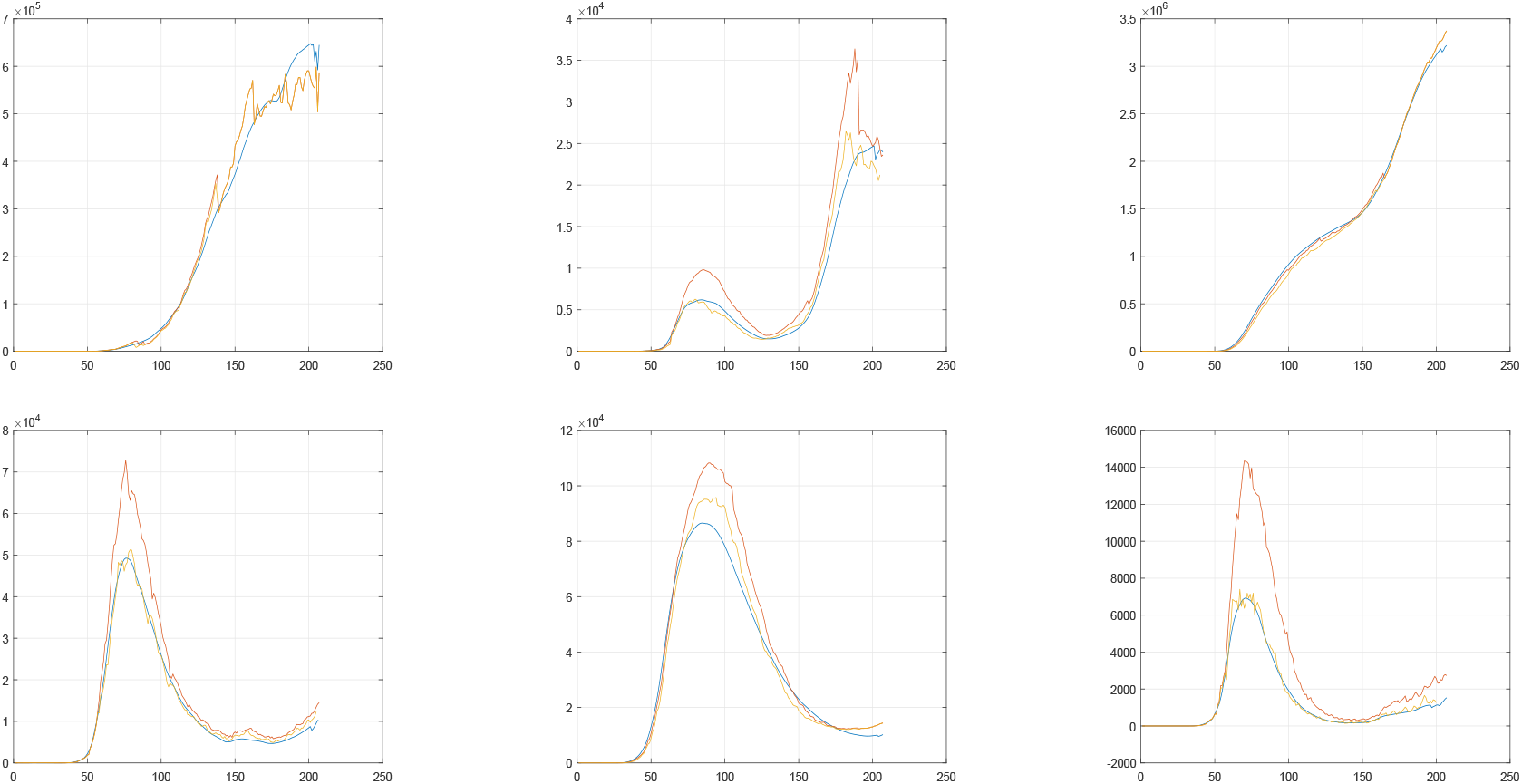
Number of infected cases. Red: raw data, yellow: inferred from shifted raw data, blue (working version): regression-adjusted. From top left to bottom right: Brazil, Israel, USA, Germany, Italy, Switzerland.

But once it is realized that shifting in time the number of recovered cases markedly improves the SIR *γ*-equation fit, perhaps it is possible to attempt a gradual shift in time, where the recording of recovered cases turns progressively more efficient.

**Sharp jumps of the number *RC* of recovered cases**. Suppose that on a particular day the number of newly recovered cases is five times the daily average in the next week. It makes sense to shift the number of recovered cases (by four or less days) backwards in time before this date rather than globally. This procedure is performed in small steps, as follows. Let ∆*RC*_1_(*t*) be the number of new recovered cases in day *t* and let ∆*RC*_2_(*t*) be the daily average of new recovered cases in the week following day *t*. Suppose that the ratio 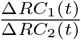, taken from day 40 or 50 to a week before the end of observation, is maximal at day *t*_1_, where it exceeds some threshold like *TH* = 3. The function *RC*(*t*) will be modified as follows: It stays as is from day *t*_1_ onwards, it is shifted backwards by one day before day *t*_1_, and the single day left unassigned is given as value the simple average of the two days next to it. The procedure is applied again and again to the newly defined function, until no ratio exceeds the threshold, or some maximal number of steps, such as 5 or 10, has been performed. The rationale behind choice of *TH* is that after adding a day when the ratio is *TH*, the new ratio will be 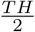. Hence, *TH* should be somewhere between 2.5 and 3. The choice *TH* =2.5 is adopted.

If the maximal allowed number of steps is kept small, the modified vector of numbers of recovered cases can be then submitted to the global DAY procedure above to account for a possible delay towards the end of the observation period.

The output of this delay procedure, with the original empirical cumulative number of affected and dead cases and the modified number of recovered cases, satisfying the SIR *γ*-equation, should be the input for Section 4, dealing with the SIR *β*-equation: estimation of *α* and *β* via flexible *K*. However, the estimates of *γ* reported in Table 1 are too different. A sanity check is called for. This is the subject of the next section.

**Figure 3:**
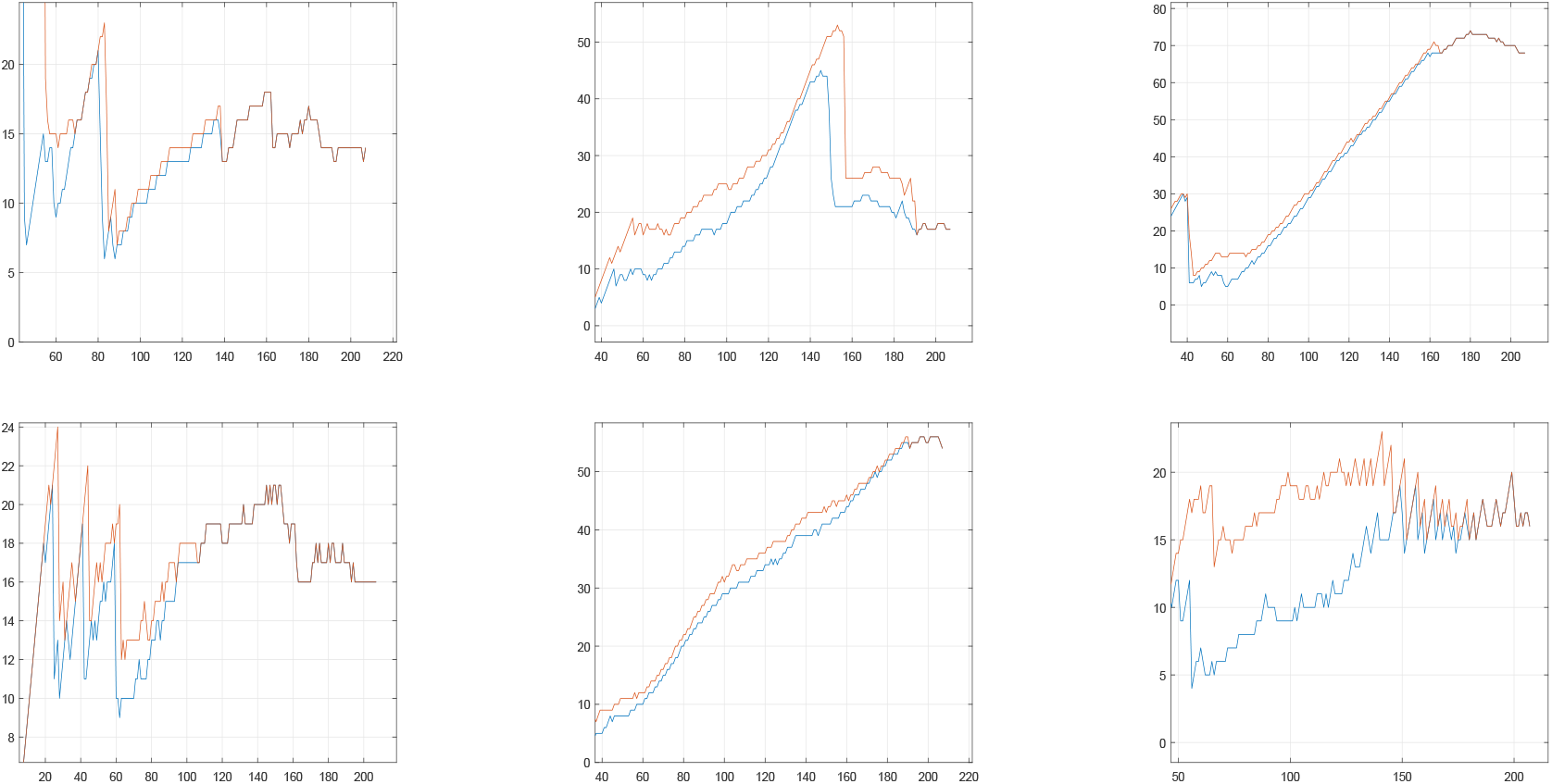
Horizontal distance between the affected and removed curves. Red: raw removed data. Blue: shifted removed data. From top left to bottom right: Brazil (stable around 15), Israel (stable around 18-20, singularity between the twowaves), USA (under-reported recovered cases), Germany (stable around 16), Italy (under-reported recovered cases) and Switzerland (stable around 10-15).

## 3 Instability of *γ*: the deficient report of recovered cases

The meaning of *γ* is perhaps carried more crisply by 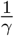, the theoretical average time an infected individual spends as infected until removed (recovered or dead). Table 1 reports values too different for *γ* in the various countries illustrated. It is inconceivable that this average could be 100 days in USA and 27 in Italy on the one hand, and between 8 and 16 days in the other four countries. The ill fit of Italy in Figure 2 after the regression correction indicates that something is not right. The horizontal distance between the affected and removed curves in Figure 8 should in principle be this average infection time 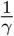, the time it takes from the moment the number of affected cases reaches some height until an equal number of cases has been removed.

Figure 3 displays these horizontal distances. In a nutshell, the horizontal distance is roughly 16 days. In periods between waves of the epidemic, when the number of affected cases stays roughly constant, this distance is ill defined, with a tendency to appear bigger. This is apparent in Germany, Israel and Switzerland. Brazil never stabilized, and the constancy of the horizontal distance is apparent. Except for the between-waves instability, all four countries conform with the 16-day statement.

Italy and the USA are different. The horizontal distance is a steadily increasing function. The proposed explanation is that in these two countries the recovered cases are only partially reported, and the Data Repository data is defective.

It is clear that data omission in the USA far exceeds that in Italy. There are states of the US, such as California and Florida, for which the entire column of recovered cases is identically zero. Correspondence with CSSE at Johns Hopkins University has corroborated awareness of this phenomenon. A warning message is attached as an Appendix.

The remainder of this section is dedicated to an exercise. The parameter *γ* is fixed as 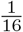 in all countries illustrated, the data on recovered cases is ignored, and the number of infected cases *Ŷ* is derived from the numbers of affected and dead cases only, by means of formula (7). Figure 10 displays a comparison with Figure 9, where each country is assigned its estimated *γ*. Equivalently, Table 2 presents parameter estimation under 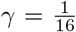, compared with Table 1, where each country is assigned its estimated *γ*. Due to the observed under-reporting of recovered cases, the number of infected cases in the USA, and to a lesser extent also in Italy, is markedly smaller in the more realistic Figure 10 than in the raw-recovered-data Figure 9.

As the purpose of the current “tutorial of sorts” is to communicate a personal outlook on data exploration rather than presenting a country-by-country analysis as such, this exercise is presented and then ignored for the rest of the paper.

## 4 Affected cases -estimation of *α*, *β* and *K*

*This Section identifies an interval of time on which epidemic dynamics are as stationary as feasible, to apply as the training period. On this period, the target population size K is viewed as a parameter, estimated together with α and β, so as to satisfy the SIR β-equation as closely as possible*.

The SIR *β*-equation (3) may be expressed for fixed *K* as

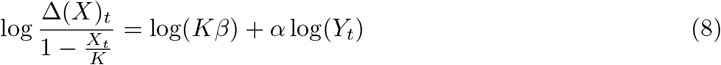

For given *K*, this is a regression equation with slope *α* and intercept 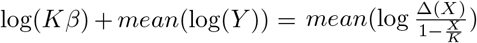, from which *β* can be derived as a function of *α*.

**Figure 4:**
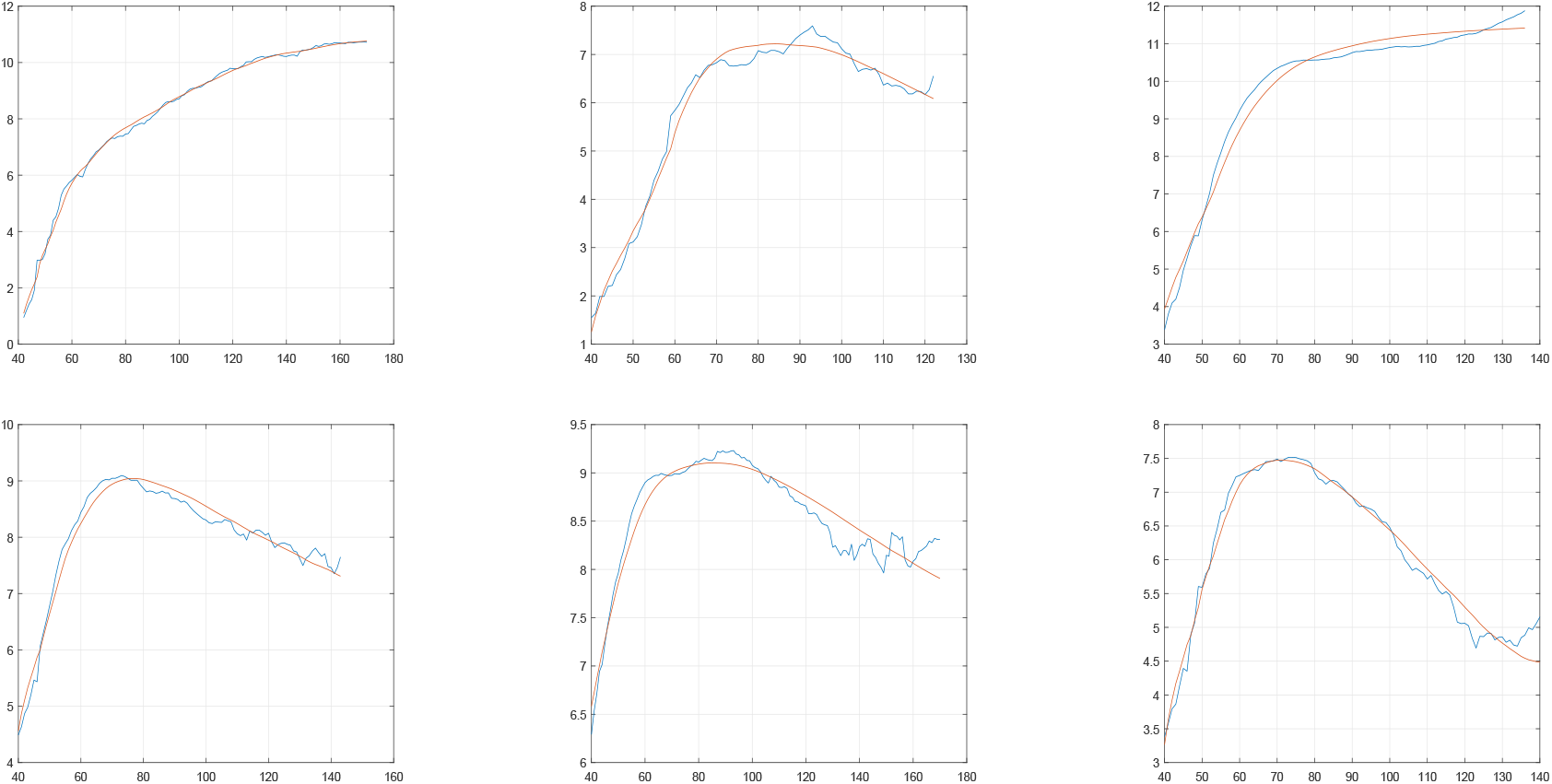
Logarithmic incremental affected cases (blue) and their linear regressions on logarithmic infected cases (red), each country in its training period. From top left to bottom right: Brazil, Israel, USA, Germany, Italy, Switzerland.

**The training period** [*M*_1_*,M*_2_]. The first task is to identify a period of stationarity, to be applied as the training period. It starts not earlier than *T*_1_, say 40, and ends not earlier than *T*_2_, say 90, nor later than *T*_3_, say 170. If there are indications that the first two constraints are too wide, the country is left without analysis. Attempts are made to base the definition of the training period on the relatively smooth function *Y* only.

*M*_1_ - **beginning of the training period**. Rather arbitrarily, the training period will be set to start at the first day *M*_1_ from day *T*_1_ onwards, when the number of infected cases *Y* is at least some number, like 5.

**Table 1:** Parameter estimation, recovered cases calibration and determination of training period.

| Country | $\alpha$ | SE $\alpha$ | $K\beta$ | $\gamma$ | $K$ | $M_1$ | $M_2$ | Delay |
| --- | --- | --- | --- | --- | --- | --- | --- | --- |
| Brazil | 0.8323 | 0.0737 | 0.8282 | 0.0643 | 9.6 M | 42 | 170 | 3 |
| Israel | 0.9141 | 0.1019 | 0.4675 | 0.0555 | 17100 | 40 | 122 | 11 |
| USA | 0.7516 | 0.0775 | 2.2773 | 0.0099 | 2.24 M | 40 | 136 | 10 |
| Germany | 0.7680 | 0.0768 | 2.1045 | 0.0841 | 218300 | 40 | 143 | 5 |
| Italy | 0.6984 | 0.0605 | 5.0417 | 0.0370 | 255000 | 40 | 170 | 5 |
| Switzerland | 0.8058 | 0.0860 | 1.4178 | 0.1242 | 35500 | 40 | 140 | 8 |

*M*_2_ - **end of the training period**.

The first date to identify is the day *t*_1_ when 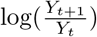 is minimal. Let *LRY*_1_ be this minimal value.

**Table 2:**
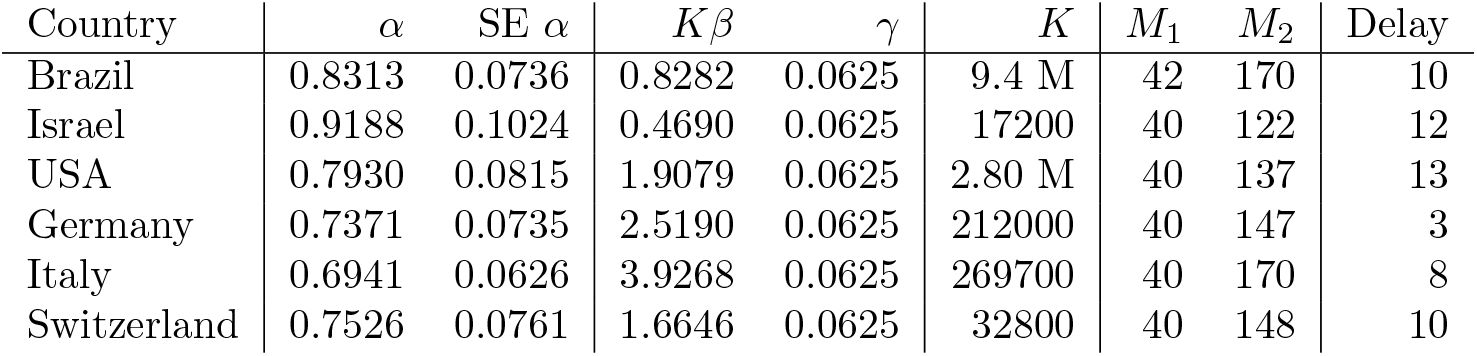
Parameter estimation (other than *γ*, fixed at 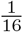), recovered cases calibration and determination of training period.

If *LRY*_1_ ≥ 0, *M*_2_ is taken as *t*_1_, where the increasing function log(*Y*) turned locally from concave to convex.

If *LRY*_1_ *<* 0, identify next the maximal date *t*_2_ such that *Y* is an increasing function on [*M*1*, t*_2_], and let *LRY*_2_ be the minimum of 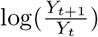 on [*t*_2_*, T*_3_], attained at day *t*_3_. If *t*_3_ = *T*_3_*,M*_2_ = *T*_3_. This means that *Y* increased until day *t*_2_ and log(*Y*) oscillated thereafter without experiencing a significant local drop.

It remains to define *M*_2_ if the significant drop occurred at *t*_3_ *<T*_3_. In this case, *M*_2_ is the maximal *t* ≤ *T*_3_ such that *Y* is decreasing on [*t*_3_*,t*].

**Solving the regression equation (3)**. This is a regular regression equation on [*M*_1_*,M*_2_] for fixed *K*. *K* is chosen by maximizing the correlation coefficient between the independent and dependent variables. Figure 4 displays the dependent variable and its linear predictor.

There are alternative estimation methods. Alon and Meilijson [1] adopted the random time transformation method of Bassan et al [2] for parameter estimation in differential equations, similar to the diffusion methods via Fokker-Planck equations. Whichever method is used, the triple (*α, β, γ*) can be then substituted in the ISPS formula (3).

## 5 Country by country results

The maximal number of backward shifts of recovered cases allowed was 10, except for Switzerland where it was taken as 1 because of numerical instability. The global DAY delay added six more days.

Table 1 presents the results of the statistical analysis of the countries under study.

Brazil and Switzerland have *α* at distance 2.3 SE below 1. For Israel the distance is 0.84 SE, for Germany and USA 3 SE and for Italy 5 SE. The parameter *α* is thus significantly fractional.

If USA had consistently twice the number of recovered cases, its parameter *β* would have been closer but still smaller than that of other countries. Its proportion of dead cases would then compare with those of Brazil and Italy, with all three stabilizing towards the end of the observation period at twice those of Germany, Israel and Switzerland.

## 6 ISPS -Implied susceptible population size

This notion, defined in (3) and explained in the Introduction, mimics implied volatility in Finance.

Figure 5 displays ISPS during the observation period and its projection into the near future, by the scenario to be described in Section 8).

Ignoring dates beyond the end of the observation period (August 15, day 207), it is possible to see that of the six countries, Brazil stands out in the stability of *K* around 9 million, with ISPS only slightly above it towards the end of the observation period. All other five countries display to a larger or smaller extent stability of *K* between days 90 and 120, and increasing ISPS thereafter. The epidemic appears as gradually affecting an expanding target population.

**Figure 5:**
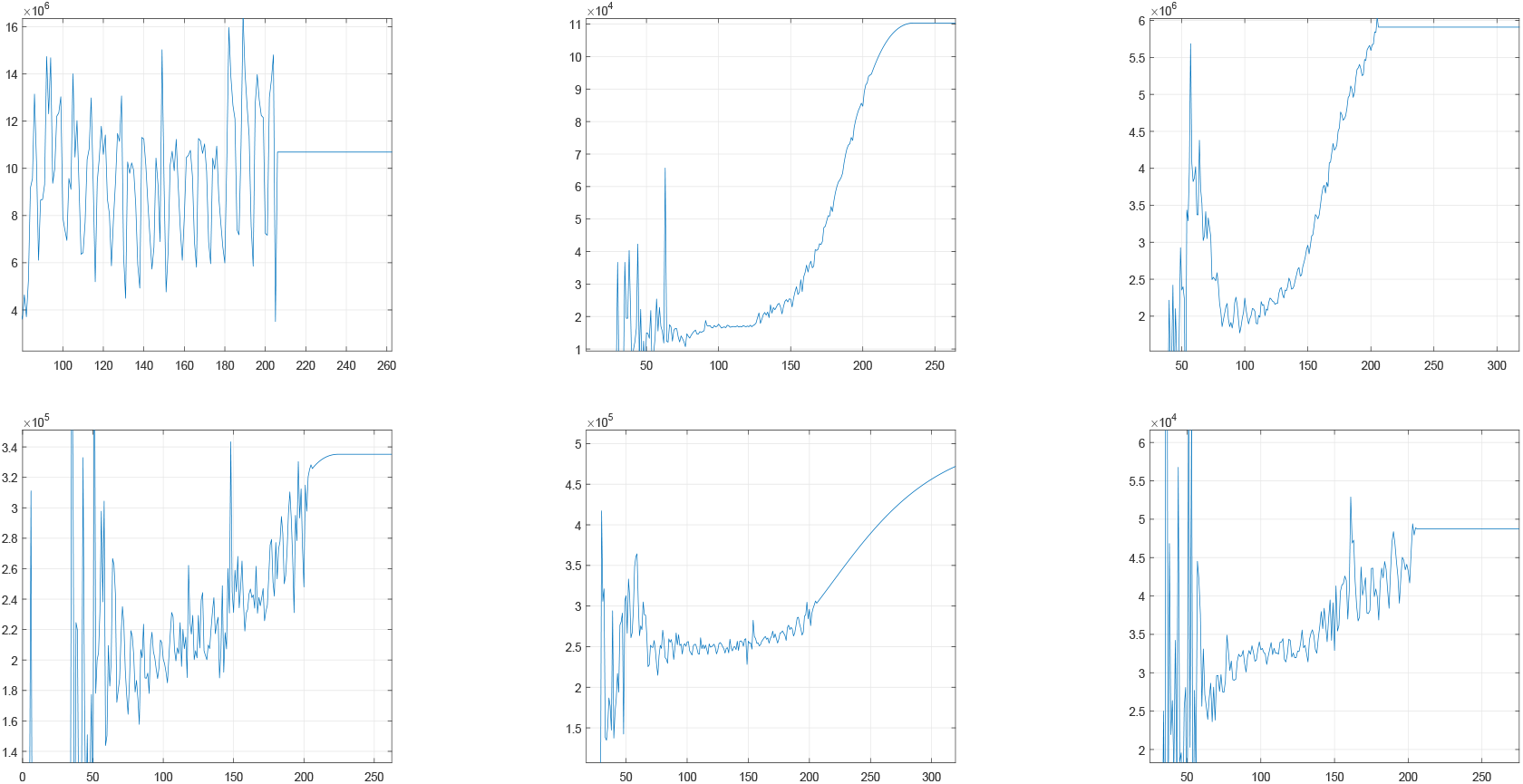
ISPS: Implied susceptible population size ISPS. Empirical measurement until day 207 (August 15) and projected scenario beyond. From top left to bottom right: Brazil, Israel, USA, Germany, Italy, Switzerland.

Since (the ideally constant) ISPS function is viewed as a stochastic process, it is convenient to monitor its logarithmic daily rate of growth function “Delta Log K” 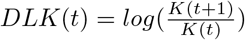. Under stationary behavior, this function should in principle be zero and in practice it should oscillate around zero. It should be stressed that a period of (positive) constancy of DLK signifies exponential growth of the target population. Let us examine DLK in greater detail.

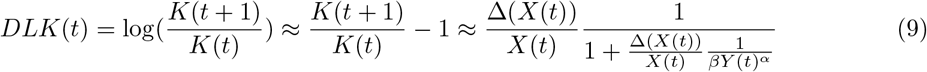

Approximation (9) suggests that *DLK* should oscillate markedly for small *t* and barely oscillate for large *t*. This is indeed the case, as would have been seen if Figure 7 extended back in time.

On the other hand, *XDLK*(*t*)= *X*(*t*)*DLK*(*t*) should be nearly homoscedastic. The more regular behavior of *XDLK* lends itself for fitting a trend curve for the purpose of setting a reliable estimate of *DLK* towards the end of the observation period. The chosen tool has been linear regression of *XDLK* on time during the last month of the observation period, to avoid giving undue weight to non smoothed data. These linear functions were submitted to “testimation”. If the slope is significant, the linear fit on XDLK is adopted and translated to *DLK*. If the slope is not significant but the intercept is, the constant fit is adopted. If none is significant, *DLK* is deemed zero.

This procedure was not applied to modify the observed data. It was performed for the sole purpose of building future scenarios, as described in Section 8.

## 7 Removed cases divided into recovered and dead cases

This division is not part of the standard SIR equations. However, it is important on its own sake, and can shed some light on data quality. Figure 6 displays the proportion of incremental dead cases out of incremental removed cases, smoothed by a two-week-window moving average. As a rule, all countries displayed show a sharp decrease of this proportion along time. USA shows a decrease from 40% to 3.5% with *γ* estimated as 0.01. The parallel decrease in Germany and Switzerland is from 4% or 5% to less than 1%, with respective *γ* estimated as 0.085 and 0.125. Israel shows a decrease from 2.5% to less than 1%, with *γ* estimated as 0.056. Brazil and Italy stand in between, with *γ* assessed respectively as 0.064 and 0.037 and lethality proportion decreasing from 10%-20% to 2%.

This disparity may suggest that the identification of Covid19 as infection and cause of death differ markedly between countries. This issue needs attention.

**Figure 6:**
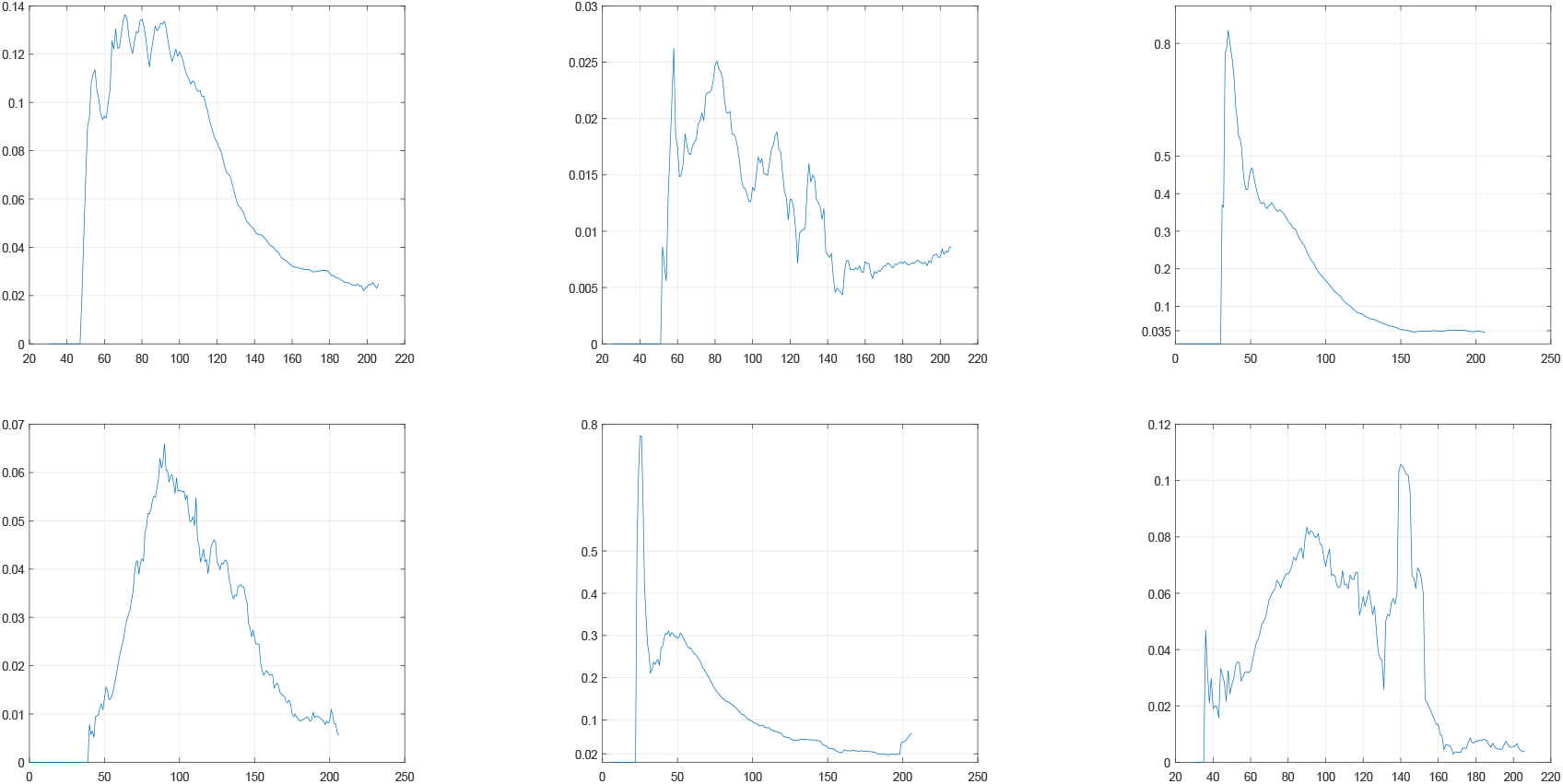
The proportion of incremental dead cases out of incremental removed cases. From top left to bottom right: Brazil, Israel, USA, Germany, Italy, Switzerland.

## 8 Scenario analysis -ISPS beyond the observation period

The plan is to extend the process *DLK* into the future as a scenario. This invented projection -not prediction -is the solution to the SIR equations with the parameters *α, β, γ* fitted in the observation period, and variable *K* derived as ISPS from the DLK extension. Consider the linear function fitted to XDLK in the last month of the observation period, translated to DLK scale as the red lines in Figure 7. Replace this translation by a linear function going through the two endpoints, and extend this linear function into the future, discounted exponentially at a daily rate of 2.5%, but constrained to be non-negative. The result is the surrogate DLK function plotted in Figure 7 beyond the observation period. The corresponding ISPS functions are plotted in Figure 5, and the solutions to the SIR equations, during and beyond the observation period, are plotted in Figure 8. The projected behavior of the number of infected cases becomes apparent. The dates and heights of the local maxima are marked.

The projected numbers of affected, removed and infected cases have been plotted for two or three months beyond the observation period for illustration purposes. There is no solid basis for claiming predictive power.

**Figure 7:**
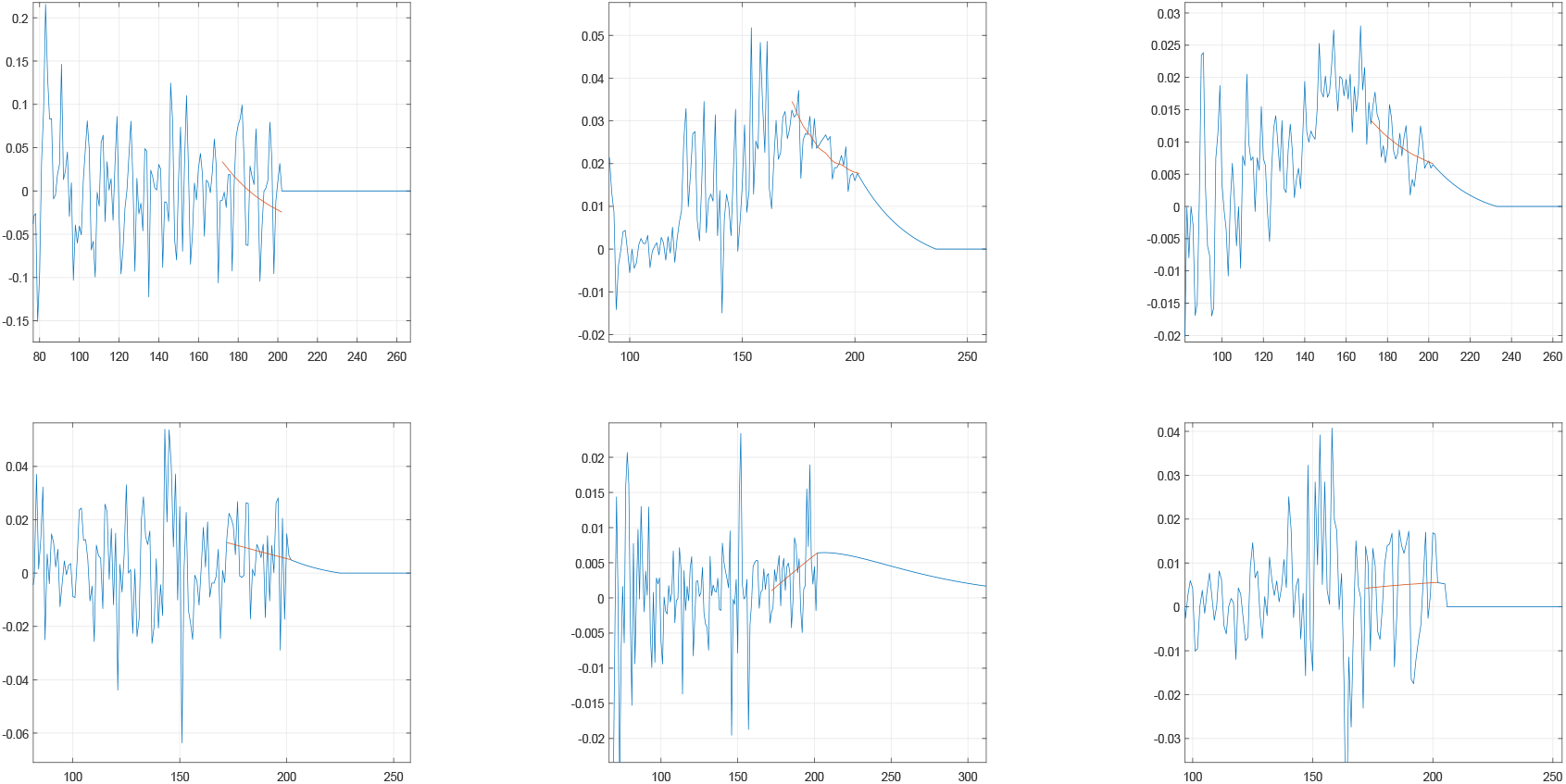
DLK: Incremental logarithmic ISPS. Empirical measurement until day 207 (August 15) and projected scenario beyond. From top left to bottom right: Brazil, Israel, USA, Germany, Italy, Switzerland.

## 9 Conclusions

Analysis of Covid19 dynamics has been performed on and restricted to the most reliable publicly available data, the data repository at Johns Hopkins University. Evidence has been found that recovered cases are only partially recorded in some countries, and the delay in recognizing recovered cases seems longer than the recognition of affection or death. Attempts to adjust the data so as to correct effects of this delay, have led to date-aligned data that satisfies the relation between removed and infected cases claimed by the SIR *γ*-equation in epidemiology (Kermack and McKendrick [7]).

The model followed is at variance with the usual interpretation of the SIR *β*-equation that relates the number of new affected cases to the number of currently infected cases and the size of the susceptible population. Evidence has been provided that a fractional power *α<* 1 of the number of infected cases (as suggested by Grenfell et. al. [4]) fits data better than the number of infected cases itself (*α* = 1), and that it is justified and useful to allow the data to uncover the extent of the susceptible population.

The size of the susceptible population has been considered a parameter to be estimated together with *α, β* and *γ* in a period of stability, and inferred on daily data thereafter, as ISPS, the implied susceptible population size. ISPS can be instrumental in extending dynamics to the near future following scenario assumptions.

The methods presented have been illustrated on the data of each of six countries.

## Data Availability

The data analyzed in this work is taken from the COVID-19 Data Repository by the Center for Systems Science and Engineering (CSSE) at Johns Hopkins University.

## Acknowledgements

Thanks are due to Nitay Alon, Yoav Benjamini, Ilan Eshel, Peter Bickel, Eytan Ruppin, Laura Sacerdote and David Steinberg for helpful suggestions.

---

**Figure 8:**
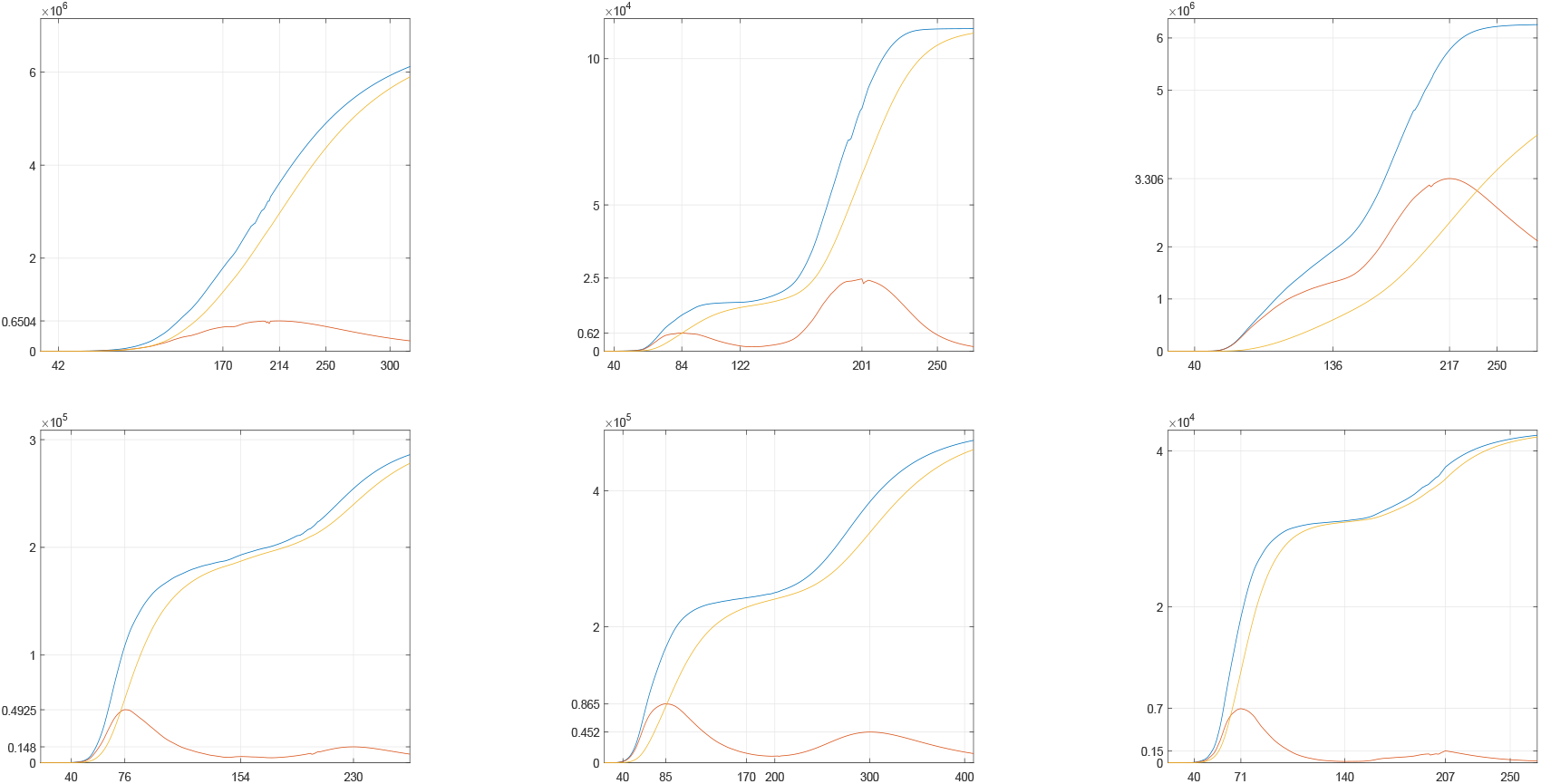
SIR equations solution under the scenario, with marked local maxima of the number of infected cases. From top left to bottom right: Brazil, Israel, USA, Germany, Italy, Switzerland.

**Figure 9:**
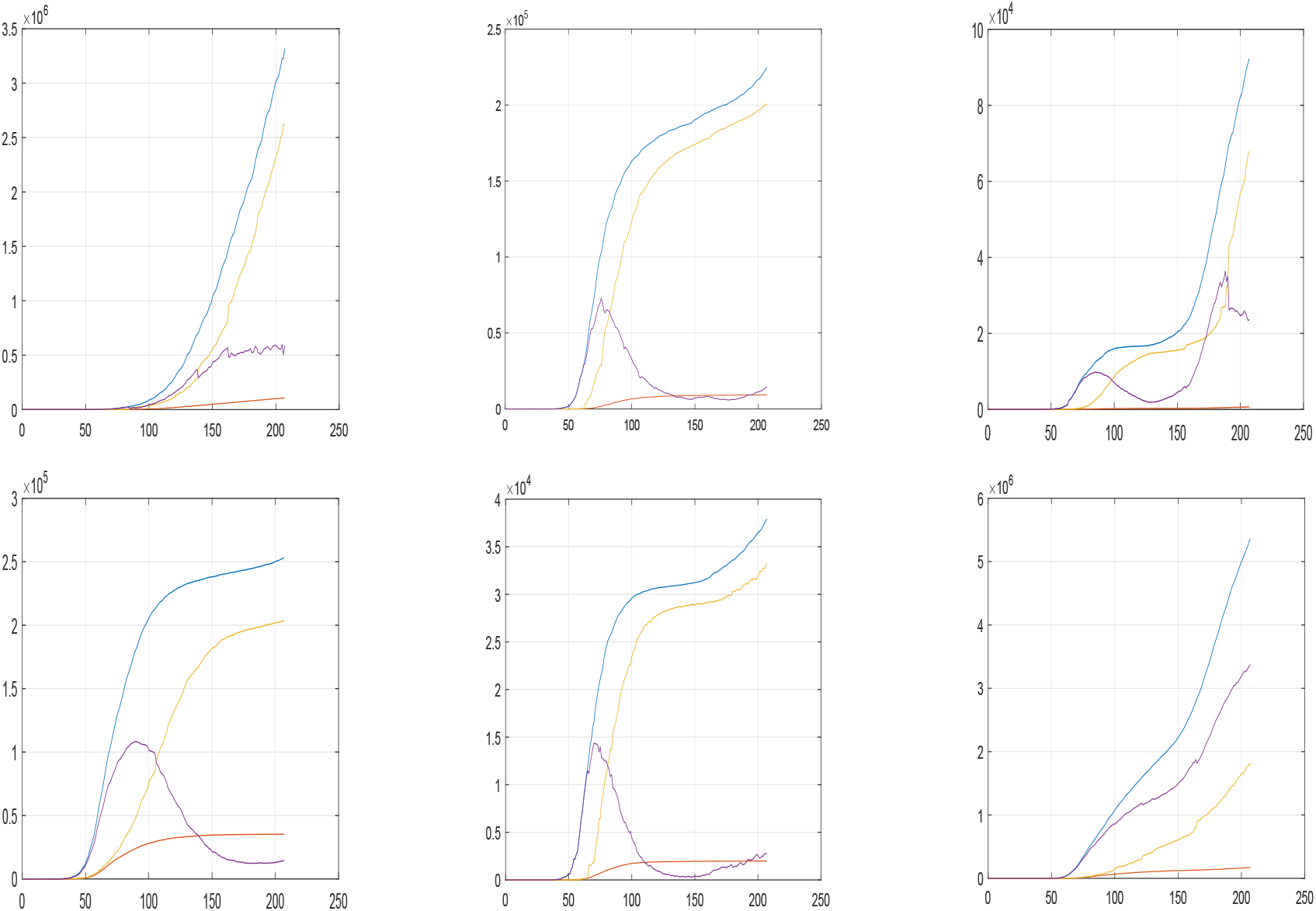
Raw Johns Hopkins data, Jan 22 2020 to August 12 2020. Blue: affected cases, yellow: recovered cases, red: dead cases, purple: infected. From top left to bottom right: Brazil, Germany, Israel, Italy, Switzerland and USA.

**Figure 10:**
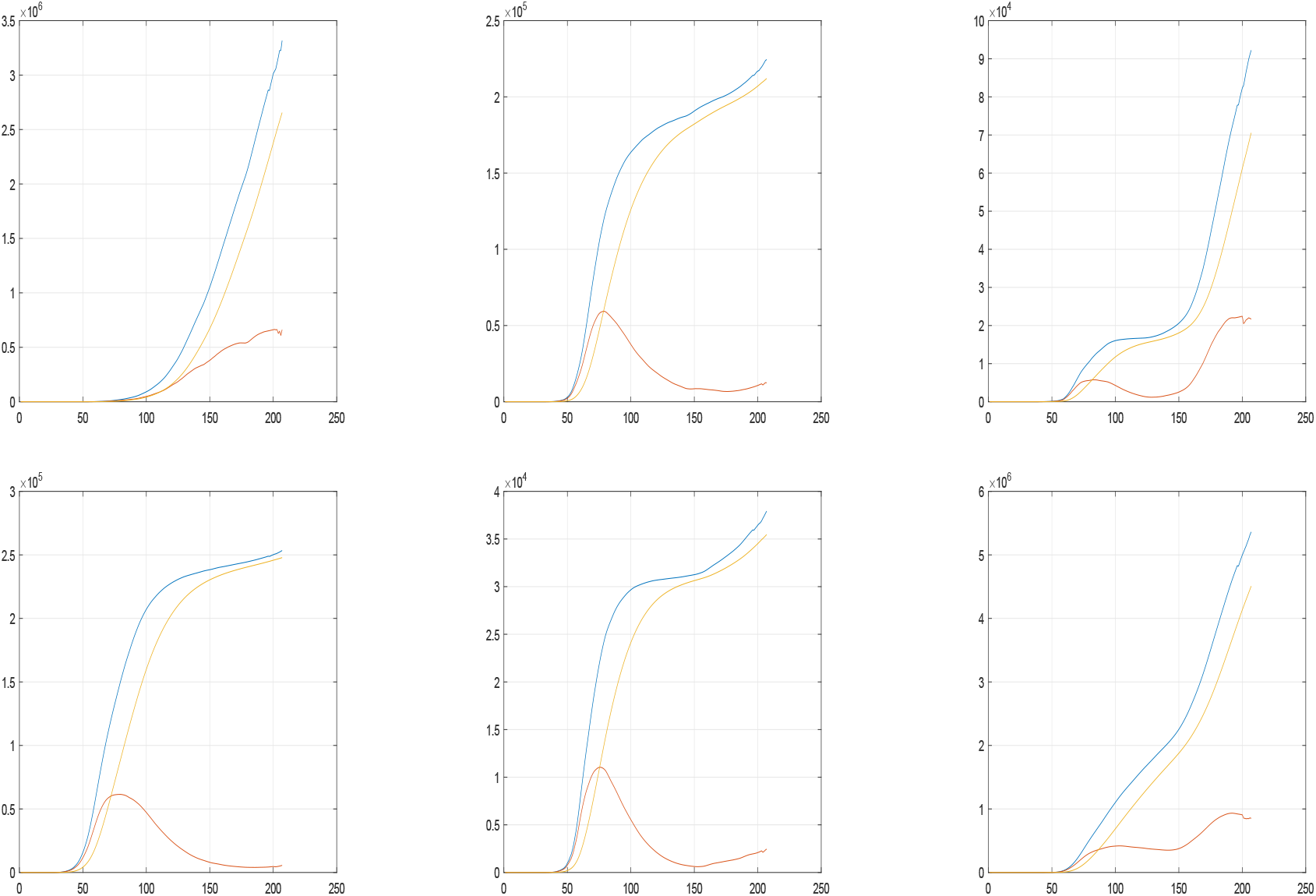
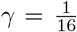 modified assessments of infected cases, Jan 22 2020 to August 12 2020. Blue: affected cases, yellow: recovered cases, red: infected. From top left to bottom right: Brazil, Germany, Israel, Italy, Switzerland and USA.

